# Implications in the quantitation of SARS-CoV2 copies in concurrent nasopharyngeal swabs, whole mouth fluid and respiratory droplets

**DOI:** 10.1101/2021.01.03.21249157

**Authors:** Priya Kannian, Bagavad Gita Jayaraman, Swarna Alamelu, Chandra Lavanya, Nagalingeswaran Kumarasamy, Gunaseelan Rajan, Kannan Ranganathan, Pasuvaraj Mahanathi, Veeraraghavan Ashwini, Stephen J. Challacombe, Jennifer Webster-Cyriaque, Newell W. Johnson

## Abstract

**Importance:** The nasopharyngeal swab (NPS) is considered the ideal diagnostic specimen for Covid-19, while WMF is recently promoted due to collection simplicity and importance in disease transmission. There is limited knowledge on the relative viral load in these samples – NPS, whole mouth fluid (WMF) and respiratory droplets (RD; another important source in transmission), on how the loads vary with disease severity and on how much virus is shed.

**Objective:** To quantify and compare SARS-CoV2 copies in the NPS, WMF and RD samples, and correlate with disease severity.

**Design:** Cross sectional study.

**Setting:** Tertiary care multi-speciality hospital with limited resources in a low-to-middle income country.

**Participants:** Eighty suspected COVID-19 patients were recruited from the COVID-19 out-patient clinic and hospital isolation wards.

**Intervention:** Concurrent NPS, WMF and RD samples were collected from all the recruited patients and tested for SARS-CoV2 copies by quantitative reverse transcriptase-polymerase chain reaction (RT-PCR).

**Main outcomes and measures:** The main outcome was COVID-19 measured by SARS-CoV2 quantitative RT-PCR in NPS samples. COVID-19 disease severity was determined according to NIH criteria. Virus shedding was defined as the presence of SARS-CoV2 copies in the WMF and RD samples.

**Results:** SARS-CoV2 was detected in 55/80 (69%) of the NPS samples. Of these 55, WMF and RD samples were positive in 44 (80%) and 17 (31%), respectively. The concordance of WMF with NPS was 84% (p=0.02). SARS-CoV2 copy numbers were comparable in the NPS (median: 8.74×10^5) and WMF (median: 3.07×10^4), but lower in RD samples (median: 3.60×10^2). Patients with mild disease had higher copies in the NPS (median: 3.46×10^6), while patients with severe disease had higher copies in the WMF (median: 1.34×10^6) and RD samples (median: 4.29×10^4). The 25-75% interquartile range of NPS SARS-CoV2 copies was significantly higher in the WMF (p=0.0001) and RD (p=0.01) positive patients.

**Conclusion and relevance:** SARS-CoV2 copies are highest in NPS samples. WMF is a reliable surrogate sample for diagnosis. High copy numbers in the NPS imply initial virological phase and higher risk of virus shedding via WMF and RD.

**Key points:** *Question:* How the numbers of SARS-CoV2 copies in nasopharyngeal swab (NPS) samples might reflectvirus shedding from the whole upper aerodigestive tract and indicatedisease severity?

*Findings:* In this cross-sectional study involving 80 suspected COVID-19 patients, the data indicate higher SARS-CoV2 copies in NPS samples of patients with mild disease,and in the whole mouth fluid (WMF) and respiratory droplet (RD) samples of patients with severe disease. Patients with higher SARS-CoV2 copies in the NPS shed the virus in the WMF and RD samples at statistically higher levels.

*Meaning:* High SARS-CoV2 copies in NPS samples imply initial virological phase withhigh levels of shedding through both WMF and RD.

## Introduction

Severe acute respiratory syndrome–coronavirus 2 (SARS-CoV2), the aetiological agent of coronavirus disease–2019 (COVID-19), is an RNA virus that infects all respiratory mucosae and the upper aerodigestive tract. Various clinical sources have been tested to choose the ideal diagnostic specimen and to help understanding of the routes of respiratory and non-respiratory transmission.^1,2^ The current gold standard for detecting SARS-CoV2 is reverse transcriptase-polymerase chain reaction (RT-PCR) using nasopharyngeal swabs (NPS).^3^ RT-PCR is a highly sensitive molecular tool that can detect very low copy numbers of the virus. Yet, this method can have low sensitivity due to inappropriate time of sample collection with regard to disease onset and diligence of sample collection in terms of appropriate trajectory reach to the nasopharynx during swab collection and adequacy of cellular material harvested.^4,5^ Recently, several studies have tested the use of saliva/whole mouth fluid (WMF) as a diagnostic specimen for the detection of SARS-CoV2 by RT-PCR, with concordance rates 80% or greater.^1,6-8^ Due to its ease of collection, WMF is being widely tested for its appropriateness as a diagnostic sample for the detection of SARS-CoV2. SARS-CoV2 has been detected in drooled unstimulated whole mouth WMF, oropharyngeal WMF and gingival crevicular fluid.^7-10^

Transmission of SARS-CoV2 is primarily through aerodigestive tract secretions including WMF, respiratory droplets (RD; >5µm particle size) and aerosols (<5µm particle size).^11^ Although many studies have shown the concordance of WMF samples for the detection of SARS-CoV2 by RT-PCR, reports on the detection of SARS-CoV2 in RD are limited to one study by Ryan *et al* and no studies have so far provided quantitative data.^12^ Previously, it has been shown with other respiratory viruses like influenza virus, that infectivity and pathogenicity is higher in droplets compared to aerosols because of the higher viral load.^13^ Therefore, quantifying SARS-CoV2 in the NPS and correlating this with disease severity and virus shedding in the WMF and RD will facilitate our understanding of disease pathogenesis.

In this study, we have validated a quantitative RT-PCR assay for the determination of SARS-CoV2 copies in the NPS, WMF and RD samples collected concurrently from suspected COVID-19 patients. We have compared the SARS-CoV2 copy numbers in the NPS with disease severity and also with the virus shedding in the WMF and RD samples. Our findings provide novel insights into our understanding of disease severity and virus transmission as well as open venues to explore methods to minimize transmission.

## Methods

### Patients and samples

The study was approved by the VHS-Institutional Ethics Committee (proposal #: VHS-IEC/60-2020). A total of 80 patients with suspected COVID-19 symptoms were recruited after written informed consent from the out-patient department and COVID isolation wards of VHS Hospital, Chennai, India. For RT-PCR, NPS, WMF and RD samples were collected concurrently from all patients in our study. The NPS samples were collected in 3ml of viral transport medium (VTM). Unstimulated WMF samples were collected in sterile wide-mouthed screw-capped containers by drooling. RD samples were collected onto Whatman No. 1 filter paper discs (diameter: 9cm; particle retention: 11µm) placed inside face masks. Only RD will be retained while aerosols would have passed through. The patients were asked to exhale deeply five times onto the paper discs, which were folded and sealed into zip-lock plastic bags. All three samples were transported to the VHS Laboratory immediately. The NPS and WMF samples were stored at 4°C and processed within 24 hours. The RD samples were stored at room temperature in a cool dry place until further processing. All SARS-CoV2 RT-PCR positive patients with COVID-19 were stratified to have mild, moderate or severe disease based on the NIH criteria.^14^

### RNA extraction and RT-PCR

The NPS samples were vortexed gently for 15 seconds. The swab was then removed and discarded. The samples were centrifuged at 3500 rpm for 10 mins. The supernatant was discarded and the cell pellet was resuspended in the bottom 750µl of the VTM. From here, 200µl was mixed with 560µl of RNA lysis buffer (QIA Amp Viral RNA kit, Qiagen, Germany) and RNA extraction performed in the automated nucleic acid extractor (QiACube Connect, Qiagen, Germany) as per manufacturer’s instructions. WMF samples were mixed with an equal volume of saline to break the mucous and centrifuged at 3500 rpm for 10 mins. The supernatant was discarded and the cell pellet resuspended in the bottom 1ml of WMF. From here, 200µl of the sample was processed as described above for NPS. For RD samples, the marked area was cut into small pieces and incubated in 700µl of RNA lysis buffer at 37°C for one hour with intermittent gentle agitation. The buffer was then collected and processed as described above for NPS. Five microliters of the eluted RNA was added to a 15µl mastermix (Labgenomics Labgun AssayPlus or Exofast, Siemens, Germany). The single step RT and amplification of *N gene* and *RdRp gene* with an internal control gene was carried out in a Lightcycler 96 (Roche, USA).

### Quantitative RT-PCR

Commercially available SARS-CoV2 RNA standards of *N gene* and *RdRp gene* (Exact Diagnostics, USA) were used to generate standard curves. The analytical sensitivity of the RT-PCR was determined using serial 10-fold dilutions of the standards in duplicates beyond the limit of detection in two independent experiments. The viral copy numbers in the clinical samples were extrapolated from the cycle threshold (Ct) values using the standard curve equation.

### Clinical sensitivity of RT-PCR

A known high positive sample was serially diluted in 10-fold dilutions using a pooled sample of five known negatives. This was done for both NPS and WMF samples. From each of these dilutions, 200µl was mixed with 560µl of RNA lysis buffer, and the same protocol as above followed. For RD, the SARS-CoV2 standard was diluted in water, impregnated onto the filter paper discs and air dried. These discs were then subjected to RNA extraction as above.

### Statistics

Mean and median were calculated using Microsoft excel. McNemar’s test, Chi-square test and t-tests were done using free online calculators from VassarStats and Social Science Statistics.

## Results

### WMF can be used as a surrogate sample for the detection of SARS-CoV2

NPS is the gold standard for the detection of SARS-CoV2 by RT-PCR. Collection of NPS requires medical expertise, and there is some discomfort to the patient. WMF is a less invasive sample that can be self-collected. Therefore, in this study we compared the sensitivity and specificity of WMF with that of the NPS samples in the detection of SARS-CoV2 by RT-PCR. Of the 80 patients recruited, SARS-CoV2 RNA was detected in 55 (69%) patients in their NPS samples. SARS-CoV2 RNA was detected in the WMF of 44/55 (80%) NPS positive patients (Table 1). Thus, the sensitivity of detecting SARS-CoV2 RNA in the WMF is 80% and the specificity is 92%. The concordance rate of 84% for the WMF samples in comparison with the NPS samples was statistically significant (p=0.02; McNemar’s test). Thus, in most cases, the easy to collect WMF samples could be used as a reliable surrogate sample for the detection of SARS-CoV2 RNA.

**Table 1:** Sensitivity and specificity of WMF samples for the detection of SARS-CoV2 by RT-PCR.

| **NPS** | **Total N (%)** | **WMF Positive** | **WMF Negative** |
| --- | --- | --- | --- |
| Positive n (%) | 55 (69) | 44 (80) | 11 (20) |
| Negative n (%) | 25 (31) | 2 (8) | 23 (92) |

True positive (TP) = 44
True negative (TN) = 23
False positive (FP) = 2
False negative (FN) = 11
Sensitivity = $TP / (TP+FN) = 44 / (44+11) = 44 / 55 = 80\%$
Specificity = $TN / (TN+FP) = 23 / (23+2) = 23 / 25 = 92\%$
Concordance = $(TP+TN) / 2 = (44 + 23) / 2 = 84\%$ (p = 0.02; McNemar's test)

### Quantitation of SARS-CoV2 RNA copies and clinical sensitivity of RT-PCR

The commercially available RT-PCR kit was validated in-house for quantitation using known SARS-CoV2 RNA standards. The in-house analytical sensitivity was determined to be 250 copies/ml, which is the same as the manufacturer states. The clinical sensitivity was determined individually for all the three sample types – NPS (305 copies/ml), WMF (345 copies/ml) and RD (453 copies/ml). Clinical sensitivity is influenced by the number of viral copies in the nasopharynx at the time of sample collection and also on adequacy of sample collection. So, we also determined the variations in the virus copies in the NPS samples collected on two consecutive days in a subset of 14 patients (Figure 1). In 11/14 (79%) patients there was a one log decrease in viral copies, while in 3/14 (21%) patients there was 2-4 log increase in viral copies: SARS-CoV2 copies may be variable in samples collected on different days.

**Figure 1:**
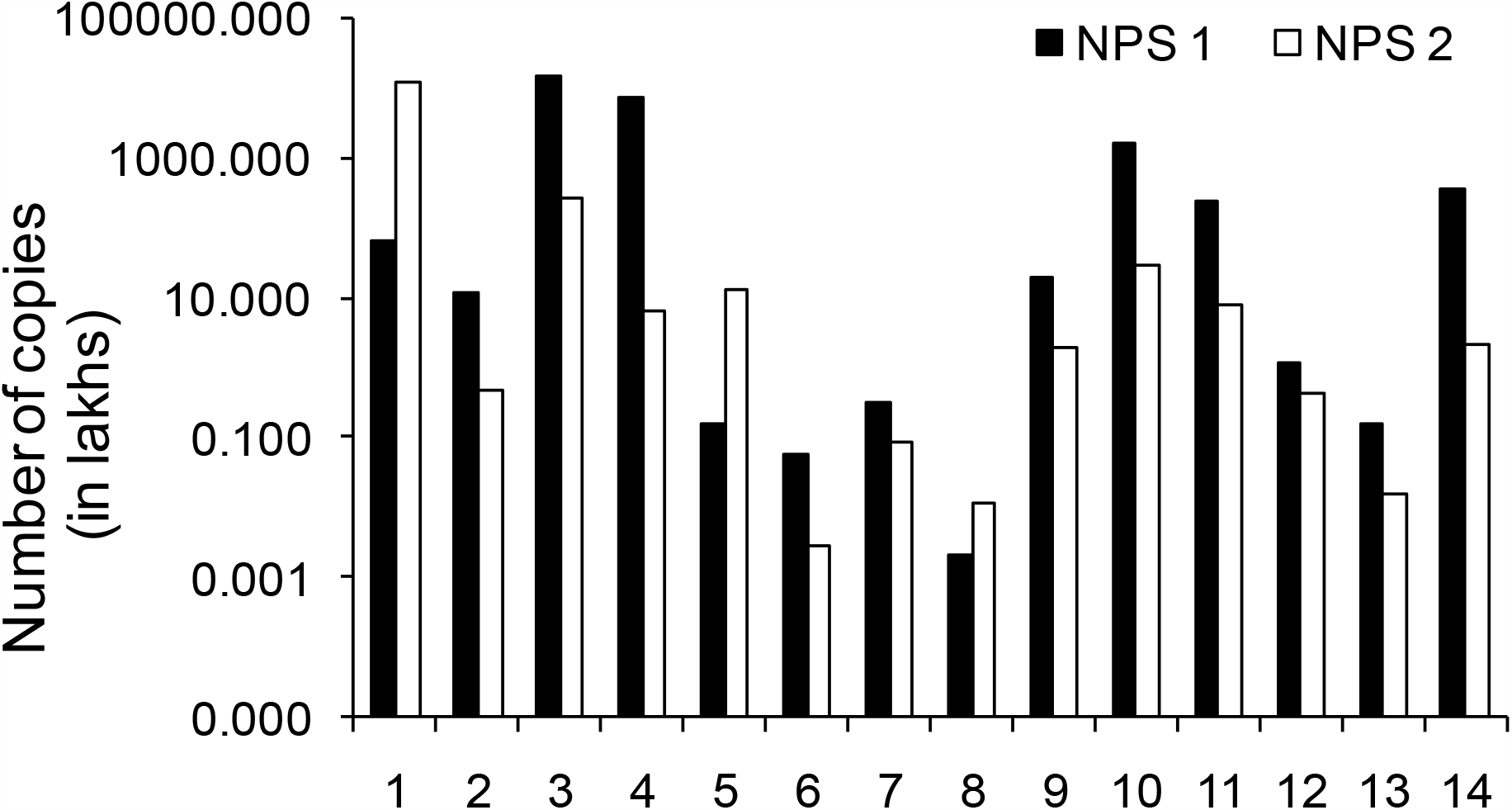
Nasopharyngeal swabs (NPS) samples collected on two consecutive days show highly variable SARS-CoV2 copies. X-axis denotes the 14 patients from whom the NPS samples were collected on two consecutive days. Y-axis denotes the number of viral copies in logarithmic scale. The black bars denote the NPS samples collected on day 1. The white bars denote the NPS samples collected on day 2 for the same patients.

### SARS-CoV2 copies are comparable in NPS and WMF samples

SARS-CoV2 copies in the NPS and WMF samples collected simultaneously in 44 positive patients were compared (Figure 2A). The median virus copies were one log higher in the NPS samples compared to the WMF samples. We also analysed the detection rates and median SARS-CoV2 copies in the NPS and WMF samples among both outpatient and inpatient cases, by gender, age and disease severity (Table 2). Patients with mild disease had higher copies in the NPS, while those with severe disease had higher copies in the WMF. However, these differences were not statistically significant. Thus SARS-CoV2 copies were comparable in both the NPS and WMF samples.

**Table 2:** Detection rates and copy numbers of SARS-CoV2 in nasopharyngeal swab, WMF and respiratory droplet samples.

| **Details** | **NPS** | **WMF** | **RD** |
| --- | --- | --- | --- |
| RT-PCR positive, n (%) | 55 (100) | 44 (80) | 17 (31) |
| Median | $8.02 \times 10^4$ | $3.07 \times 10^4$ | $3.60 \times 10^2$ |
| Out patients, n (%) | 10 (18) | 7 (16) | 2 (12) |
| Median | $1.71 \times 10^5$ | $1.32 \times 10^4$ | $1.24 \times 10^5$ |
| In-patients, n (%) | 45 (82) | 37 (84) | 15 (88) |
| Median | $8.02 \times 10^4$ | $3.25 \times 10^4$ | $2.21 \times 10^2$ |
| <u>Gender</u> |  |  |  |
| Male, n (%) | 34 (62) | 28 (64) | 9 (53) |
| Median | $4.02 \times 10^4$ | $4.54 \times 10^4$ | $3.76 \times 10^2$ |
| Female, n (%) | 21 (38) | 16 (29) | 8 (47) |
| Median | $2.36 \times 10^5$ | $9.01 \times 10^3$ | $2.69 \times 10^2$ |
| <u>Age</u> |  |  |  |
| 20-40 years, n (%) | 17 (31) | 14 (32) | 6 (35) |
| Median | $1.17 \times 10^6$ | $1.97 \times 10^4$ | $2.26 \times 10^3$ |
| 41-60 years, n (%) | 21 (38) | 15 (34) | 6 (35) |
| Median | $4.36 \times 10^4$ | $3.61 \times 10^4$ | $5.56 \times 10^2$ |
| ≥ 61 years, n (%) | 17 (31) | 15 (34) | 5 (30) |
| Median | $3.68 \times 10^4$ | $2.89 \times 10^4$ | $1.95 \times 10^2$ |
| <u>Severity</u> |  |  |  |
| Mild, n (%) | 16 (29) | 12 (27) | 5 (29) |
| Median | $3.46 \times 10^6$ | $2.28 \times 10^4$ | $8.91 \times 10^2$ |
| Moderate, n (%) | 33 (60) | 28 (64) | 10 (59) |
| Median | $8.02 \times 10^4$ | $2.42 \times 10^4$ | $2.90 \times 10^2$ |
| Severe, n (%) | 6 (11) | 4 (9) | 2 (12) |
| Median | $1.80 \times 10^4$ | $1.34 \times 10^6$ | $4.29 \times 10^4$ |

**Figure 2:**
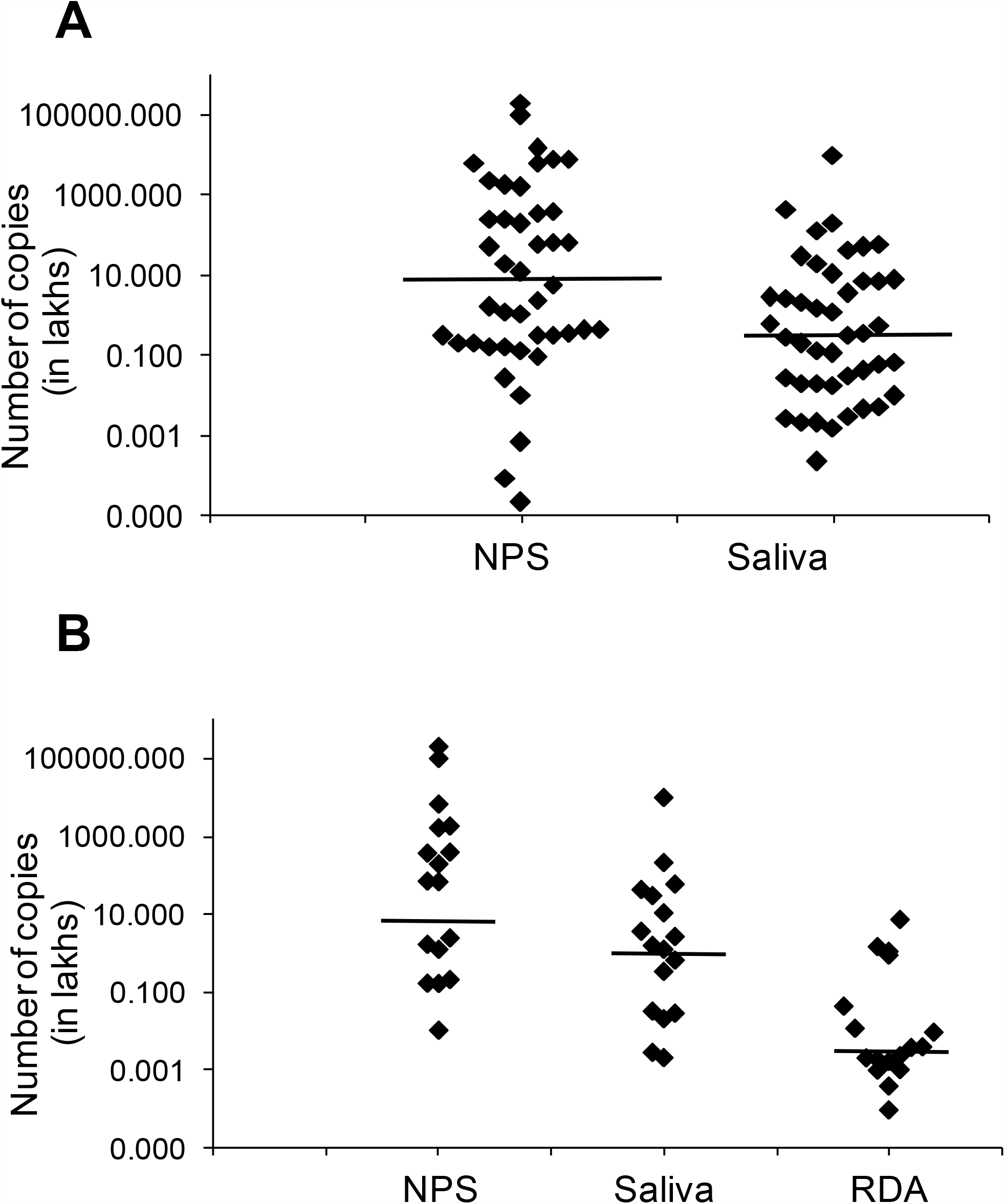
SARS-CoV2 copies in the nasopharyngeal swabs (NPS), WMF and respiratory droplets (RD) samples. A. Virus copies in the 44 patients with both NPS and WMF positive samples. X-axis denotes the sample types – NPS and WMF. Y-axis denotes the number of viral copies in logarithmic scale. The diamonds represent the samples. The short bars depict the median viral copies for each sample group – NPS: 8.74×10^5; WMF: 3.07×10^4. **B. Virus copies in the 17 patients with NPS, WMF and RD positive samples**. X-axis denotes the sample types – NPS, WMF and RD. Y-axis denotes the number of viral copies in logarithmic scale. The diamonds represent the samples. The short bars depict the median viral copies for each sample group – NPS: 6.82×10^6; WMF: 1.52×10^5; RD: 3.60×10^2.

### Detection rates of SARS-CoV2 were higher in the NPS and WMF samples compared to RD samples

We compared SARS-CoV2 copies in 17/55 (31%) patients with RT-PCR positive NPS, WMF and RD samples that were collected concurrently. The median virus copies were highest in the NPS samples and lowest in the RD samples (Figure 2B). Although the detection rate was lower in the RD samples, the median SARS-CoV2 copies were not statistically different from the NPS and WMF samples (Table 2). Similar to the WMF samples, the SARS-CoV2 copies were two logs higher in the severe COVID-19 patients compared to the mild and moderate patients.

### Patients with higher SARS-CoV2 copies in the NPS are more likely to shed the virus in the WMF and RD

We next analysed the virus copies in the NPS samples of patients who had a positive or negative WMF sample collected simultaneously. The median virus copies were three logs higher in the NPS samples of patients with a positive WMF sample (median – 8.74×10^5) compared to those with a negative WMF sample (median – 4.14×10^2). The 25-75% interquartile range was significantly higher in the patients with a positive WMF sample (Figure 3A). This difference was statistically significant (p=0.0001; Mann Whitney U test). Similarly, patients with a positive RD sample had higher median and interquartile range for the SARS-CoV2 copy numbers in the NPS samples than the negative group (Figure 3B). This difference was also statistically significant (p=0.01; Mann Whitney U test). Taken together, the data suggests that the patients with higher SARS-CoV2 copies in the NPS samples are more likely to shed the virus in the WMF and RD samples.

**Figure 3:**
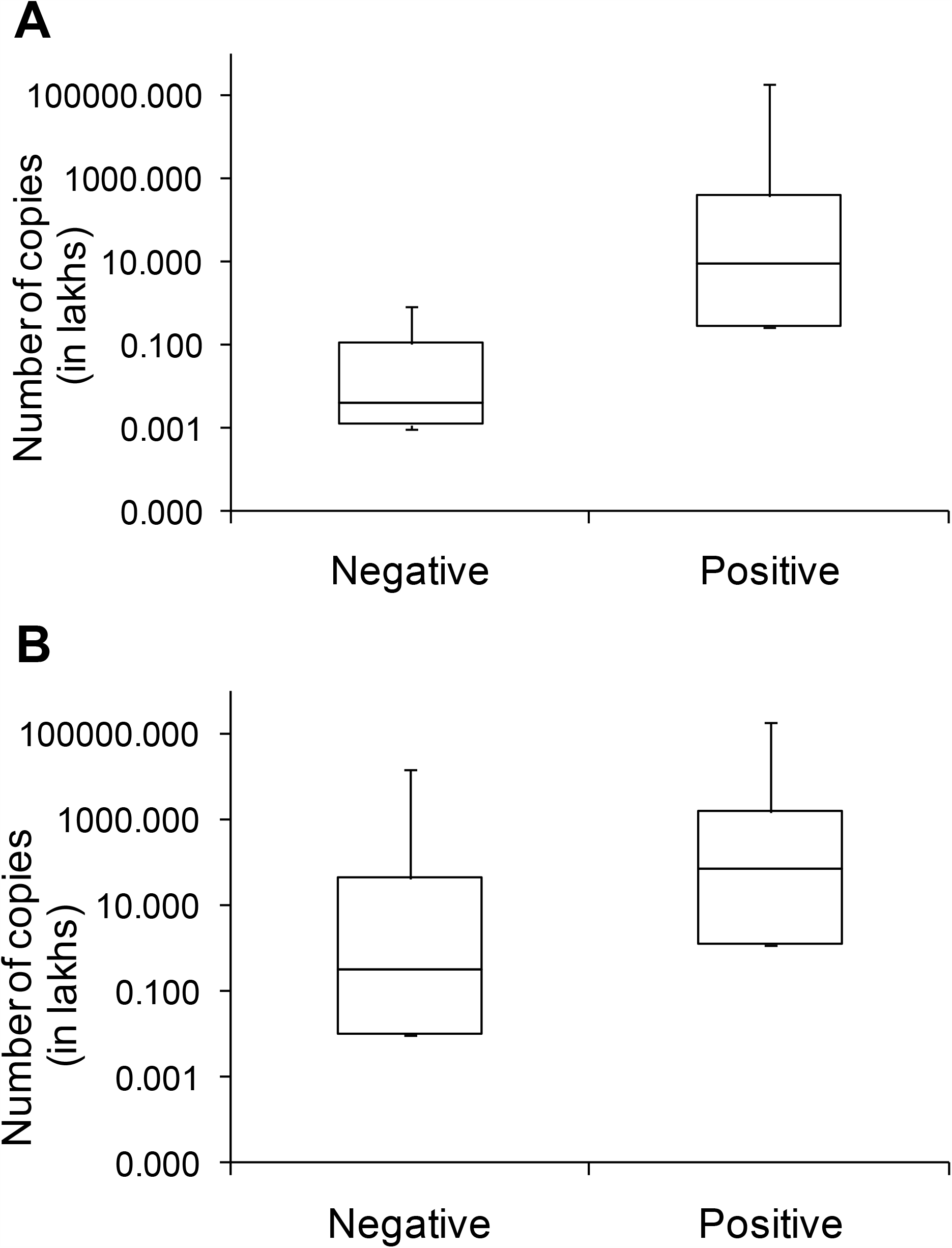
SARS-CoV2 copies in the nasopharyngeal swab (NPS) samples are higher in patients with positive WMF or respiratory droplets (RD) samples. X-axis denotes negative and positive categories of the WMF or RD samples. Y-axis denotes the number of SARS-CoV2 copies. The interquartile range shows the 25-75% range of the virus copies in each category. The error bars depict the minimum and maximum copy numbers in each category. **A**. WMF **and B. Respiratory droplets (RD)**.

## Discussion

SARS-CoV2, a respiratory RNA virus, is transmitted through respiratory secretions that are dispersed within close contacts. Initially, during the virological phase, the virus perpetuates in respiratory epithelia and elicits a host immune response. As the disease progresses from mild to severe, the host immune response takes over and produces a cytokine storm. This is the immunological phase. At the time of diagnosis, SARS-CoV2 is routinely detected by a qualitative RT-PCR method from NPS. Recently, WMF is advocated as a less invasive and easy to collect sample for RT-PCR. However, the clinical implications of carrying higher SARS-CoV2 copies are poorly understood. In this study, we have shown that SARS-CoV2 RNA copies are highest in the NPS samples followed by WMF and then RD samples. Clinical sensitivity of RT-PCR in detecting SARS-CoV2 in the NPS samples was as low as 305 copies/ml, yet the difference in the shedding of the virus between two consecutive days due to the natural course of the disease or due to variations in sample collection can cause significant variations in the clinical sensitivity of the RT-PCR in detecting the virus. Additionally, we showed that patients with higher SARS-CoV2 copies had milder disease and were more likely to shed the virus in the WMF or RD. This may be because these patients have presented relatively early in the natural history of their Covid-19 disease.

SARS-CoV2 primarily infects epithelial cells lining the oral, oropharyngeal and respiratory mucosae including endothelial cells in the lower respiratory tract and alveoli. WMF – better called WMF because it also contains serum components from gingival crevicular fluid and any mucosal inflammatory exudate - provides a more comprehensive and consistent sample than the NPS. It is a non-invasive sample that can be collected with no personal discomfort. WMF has been shown to be a good diagnostic sample in other respiratory virus infections like influenza and respiratory syncytial virus infections.^15^ Our findings show that WMF has 80% sensitivity, 92% specificity and 84% concordance with NPS in the detection of SARS-CoV2. These results are similar to other recent studies showing detection / concordance rates ranging from 83% to 91.7%.^4,7,9^ Our study has also shown that the SARS-CoV2 copies in the WMF were not significantly different from the NPS samples. Thus, WMF can be used as a surrogate sample for the screening of large numbers of people, like in rural areas where medical expertise to collect NPS samples is minimal. The small group of WMF-negative people may be confirmed by collecting NPS samples. Limitations in the use of WMF/WMF include the time to collect a diligently drooled sample, which takes about 4-5 minutes: studies on comparable performance of a quick saline mouth rinse/gargle or of stimulated WMF (example by chewing on a bland substance such as paraffin wax) are warranted.

We attempted to collect respiratory droplets from suspected COVID-19 patients simultaneously with NPS and WMF samples in a low resource setting using Whatman No. 1 filter paper discs that has particle retention of 11µm. We were able to detect SARS-CoV2 in 31% of the RD samples from NPS positive patients. Ryan *et al* showed a detection rate of 73.3% from exhaled breath condensate using two genes and 93.3% using four genes.^12^ The sample collection method (exhalation into an RTube) in Ryan *et al*’s study is superior to our resource limited collection method on a simple filter paper. Their collection time of two minutes is much longer than our five deep exhalations. These factors would contribute to the higher detection rate in Ryan *et al*’s study. In addition to detection, we have also quantitated the SARS-CoV2 copies in the RD samples: these had slightly lower median SARS-CoV2 copies, though the differences were not statistically significant.

Varying clinical presentations and virus shedding rates have been correlated with numbers of influenza virus copies in nasopharyngeal samples.^16,17^ In this study, we have shown that virus copies were about two logs higher in the NPS samples of patients with mild disease compared to those with moderate or severe disease, though this was not statistically significant. Thus, disease severity/symptomatology of COVID-19 does not correlate with SARS-CoV2 copies in NPS samples: this may reflect stage in the evolution of disease. Similarly, Lavezzo *et al* showed that there was no statistically significant difference in the virus copies between symptomatic and asymptomatic patient samples.^18^ Additionally, patients with a positive WMF and/or RD sample carried significantly higher SARS-CoV2 copies in the NPS. These findings suggest that during the initial disease onset phase, though patients have only a mild disease, they carry high virus copies, and patients with high virus copies are potent transmitters of the disease probably via WMF and/or RD. Thus, our study opens avenues for the exploration of virucidal mouthwashes or nasal sprays that could provide insights into possible ways of minimizing transmission of the virus.

## Conclusions

SARS-CoV2 detection rates and copies are highest in the NPS samples followed by WMF and RD samples. Variations in copy numbers on consecutive days throw light on the varying sensitivity of detection of SARS-CoV2 by RT-PCR. Copy numbers of SARS-CoV2 in WMF was not significantly different from those in NPS samples. This confirms that WMF/saliva is a good surrogate sample for the diagnostic detection of SARS-CoV2. High copy numbers imply mild disease or early stages in the evolution of the disease, with high potential transmission risk. It is therefore, imperative to rapidly screen and quarantine asymptomatic and mild cases.

## Data Availability

The data will not be available as a data link.

## Acknowledgements

This work was funded by intramural research funds of Chennai Dental Research Foundation, Chennai, India.

## References

1. Wang W., Xu Y., Gao R., Lu R., Han K., Wu G., Tan W. Detection of SARS-CoV-2 in different types of clinical specimens. JAMA. 2020;323(18): 1843–1844;

2. Wolfel R., Corman V.M, Guggemos W., Seilmaier M., Zange S., Muller M.A., Niemeyer D., Jones T.C., Vollmar P., Rothe C., Hoelscher M., Bleicker T., Brunink S., Schneider J., Ehmann R., Zwirglmaier K., Drosten C., Wendtner C. Virological assessment of hospitalized patients with COVID-2019. Nature 2020; 581(7809):465–469.

3. Wang H., Liu Q., Hu J., Zhou M., Yu M., Li K., Xu D., Xiao Y., Yang J., Lu Y., Wang F., Yin P, Xu S. Nasopharyngeal Swabs Are More Sensitive Than Oropharyngeal Swabs for COVID-19 Diagnosis and Monitoring the SARS-CoV-2 Load. Front. Med. (2020); 7:334

4. Williams E., Bond K., Zhang B., Putland M., Williamson D.A. WMF as a Noninvasive Specimen for Detection of SARS-CoV-2. J Clin Microbiol. 2020; 58 (8) e00776–20.

5. Higgins TS, Wu AW, Ting JY. SARS-CoV2 nasopharyngeal swab testing – false-negative results from a pervasive anatomical misconception. JAMA Otolaryngol – Head Neck Surg 2020; 146(11). doi: 10.1001/jamaoto.2020.2946.

6. To K.K., Tsang O.T., Yip C.C.Y., Chan K.H., Wu T.C., Chan J.M., Leung W.S., Chik T.S., Choi C.Y., Kandamby D.H., Lung D.C., Tam A.R., Poon R.W., Fung A.Y., Hung I.F., Cheng V.C., Chan J.F., Yuen K.Y. Consistent detection of 2019 novel coronavirus in WMF. Clin Infect Dis. 2020; 71(15): 841–843.

7. Chen JH, Yip CC, Poon RW, Chan K, Cheng VC, Hung IF, Chan JF, Yuen K, To KK. Evaluating the use of posterior oropharyngeal WMF in a point-of-care assay for the detection of SARS-CoV2. Emerg Microbes Infect 2020; 9(1):1356–1359.

8. To K.K., Lu L., Yip C.C., Poon R.W., Fung A.M., Cheng A., Lui D.H., Ho D.T., Hung I F., Chan K.H., Yuen K.Y. Additional molecular testing of WMF specimens improves the detection of respiratory viruses. Emerg Microbes Infect. 2017;6(6):e49. doi: 10.1038/emi.2017.35.

9. To K.K., Tsang O.T., Leung W.S., Tam A.R., Wu T.C., Lung D.C., Yip C.C.Y., Cai J.P., Chan J.M., Chik T.S., Lau D.P., Choi C.Y., Chen L., Chan W., Chan K.H., Ip J.D., Chin-King A., Poon R.W., Luo C.T., Cheng V.C.,Chan J.F., Hung I.F., Chen Z.,Chen H., Yuen K.Y. Temporal profiles of viral load in posterior oropharyngeal WMF samples and serum antibody responses during infection by SARS-CoV-2: an observational cohort study. Lancet Infect Dis. 2020;20(5):565–574.

10. Gupta S., Mohindra R, Chauhan PK, Singla V, Goyal K, Sahni V, Gaur R, Verma DK, Ghosh A, Soni RK, Suri V, Bhalla A, Singh MP. SARS-CoV2 detection in gingival crevicular fluid. J Dental Res 2020; 1–7. doi: 10.1177/0022034520970536journals.sagepub.com/home/jdr.

11. Siegel J.D., Rhinehart E., Jackson M., Chiarello L. Centers for Disease Control and Prevention (CDC); 2007. Guideline for Isolation Precautions: Preventing Transmission of Infectious Agents in Healthcare Settings. https://www.cdc.gov/infectioncontrol/guidelines/isolation/index.html

12. Ryan DJ, Toomey S, Madden SF, Casey M, Breathnach OS, Morris PG, Grogan L, Branagan P, Costello RW, DeBarra E, Hurley K, Gunaratnam C, McElvaney NG, O’Brien ME, Salaiman I, Morgan RK, Hennessy BT. Use of exhaled breath condensate (EBC) in the diagnosis of SARS-CoV2 (COVID-19). Thorax 2021; 76: 86–88.

13. Teunis PFM, Brienen N, Kretzchmar MEE. High infectivity and pathogenicity of influenza A virus via aerosol and droplet transmission. Epidemics 2010; 2(4): 215–222.

14. Clinical Spectrum of SARS-CoV-2 Infection. NIH COVID-19 treatment guidelines. https://www.covid19treatmentguidelines.nih.gov/overview/clinical-spectrum/

15. To K.K.W., Yip C.C.Y., Lai C.Y.W., Wong C.K.H., Ho D.T.Y., Pang P.K.P., Ng A.C.K., Leung K.H., Poon R.W.S.,Chan K.H., Cheng V.C.C., Hung I.F.N., Yuen K-Y. WMF as a diagnostic specimen for testing respiratoryvirus by a point-of-care molecular assay: a diagnostic validity study. Clin Microbiol Infect. 2019;25(3):372–378.

16. Alves VRG, Perosa AH, de Souza Luna LK, Cruz JS, Conte DD, Bellei N. Influenza A(H1N1)pdm09 infection and viral load analysis in patients with different clinical presentations. Mem Inst Oswaldo Cruz 2020; 115: e200009. doi: 10.1590/0074-02760200009.

17. To KKW, Chan K, Li IWS, Tsang T, Tse H, Chan JFW, Huang IFN, Lai S, Leung C, Kwan Y, Lau Y, Ng T, Cheng VCC, Peiris JSM, Yuen K. Viral load in patients infected with pandemic H1N1 2009 influenza A virus. J Med Virol 2010; 82: 1–7.

18. Lavezzo E., Franchin E., Ciavarella C., Cuomo-Dannenburg G., Barzon L., Vecchio C.D., Rossi L., Manganelli R., Loregian A., Navarin N., Abate D., Sciro M., Merigliano S., Canale E.D., Vanuzzo M.C., Besutti V., Saluzzo F., Onelia F., Pacenti M., Parisi S.G., Carretta G., Donato D., Flor L., Cocchio S., Masi G., Sperduti A., Catarino L., Salvador R., Nicoletti M., Caldart F., Castelli G., Nieddu E., Labella B., Fava L., Drigo M., Gaythorpe K.A.M., Imperial College COVID-19 response Team., Brazzale A.R., Toppo S., Trevisan M., Baldo V., Donnelly C.A., Ferguson N.M., Dorigati I., Crisanti A. Suppression of a SARS-CoV-2 outbreak in the Italian municipality of Vo’. Nature 2020; 584, 425–429.

